# Pharmacogenomic Variants in the Russian Population: A Retrospective Analysis of 6102 Exomes

**DOI:** 10.64898/2026.02.16.26346289

**Authors:** Anastasiia A. Buianova, Valery V. Cheranev, Anna O. Shmitko, Iuliia A. Vasiliadis, Galit A. Ilyina, Oleg N. Suchalko, Mikhail Iu. Kuznetsov, Vera A. Belova, Dmitriy O. Korostin

## Abstract

**Background:** Personalized pharmacotherapy requires systematic consideration of genetic factors influencing drug efficacy and safety. The accumulation of large-scale whole-exome sequencing (WES) data provides an opportunity to assess population frequencies of clinically significant pharmacogenetic variants; however, the diagnostic applicability of exome data for pharmacogenomics remains insufficiently studied.

**Materials and Methods:** A retrospective analysis of 6,102 anonymized sequencing datasets obtained between 2020 and 2025 was performed using the DNBSEQ-G400 (MGI) platform and Agilent SureSelect Human All Exon v6/v7/v8 enrichment kits. SNV and indel detection, CNV analysis, high-resolution *HLA* typing, and diplotype assignment for key pharmacogenes were conducted. Pharmacogenomic annotations were derived from PharmGKB (levels of evidence 1A–2B), CPIC, and PharmVar. Additionally, WES limitations and the feasibility of imputing non-coding pharmacogenetic variants were evaluated.

**Results:** Population frequencies of alleles and metabolic phenotypes were determined for 13 Very Important Pharmacogenes (VIPs), along with the distribution of *HLA* class I and II alleles. The highest allelic and phenotypic variability was observed in CYP family genes, particularly *CYP2D6, CYP2C19*, and *CYP2B6*. A total of 663 pharmacogenomic annotations were identified, predominantly related to drug metabolism (50.38%) and toxicity (29.56%), including psychotropic agents, anticoagulants, statins, opioid analgesics, antineoplastic agents, and immunosuppressants. At least 32 drugs require pharmacogenetic testing based on variants located in non-coding regions, as well as accurate *CYP2D6* copy number determination. Linkage disequilibrium analysis demonstrated the inability to reliably impute most non-coding pharmacogenetic variants from WES data.

**Conclusion:** These findings represent one of the largest reference assessments to date of pharmacogenetically significant variant and *HLA* allele frequencies in the Russian population. The results confirm the utility of WES for population pharmacogenomic screening while simultaneously highlighting its fundamental limitations and the need for alternative genetic diagnostic methods in selected cases.

## Introduction

Personalized pharmacotherapy represents one of the key directions of modern medicine, enabling the adaptation of drug selection, dosage, and treatment regimen according to the patient’s individual genetic characteristics. A substantial proportion of interindividual variability in drug efficacy and toxicity is determined by inherited variants in genes encoding drug-metabolizing enzymes (the CYP family [1], UGT [2], NAT2 [3], TPMT, NUDT15 [4]), transporters (SLCO1B1, ABCB1 [5]), drug targets, and immunogenetic markers (HLA) [6]. The economic burden associated with adverse drug reactions (ADRs) is a global concern: preventable ADRs are estimated to cost healthcare systems in the United States $2.7–5.1 billion annually £98.5 million in the United Kingdom, and approximately $42 billion worldwide, accounting for around 20 million disability-adjusted life years (DALYs) [7]. Moreover, the prevalence of ADRs is likely underestimated due to patients’ frequent failure to report them, attributable to lethargy, complacency, and ignorance [8].

An analysis of the UK national pharmacovigilance database (1963–2024) demonstrated that 9% of all reported ADRs (115,789 cases) were associated with drugs for which risk could be modified based on pharmacogenetic data. Notably, 75% of these cases were potentially preventable through testing of only three genes (*CYP2C19, CYP2D6, SLCO1B1*), with the highest proportion of such ADRs observed in psychiatric medications (47%) [9]. According to a pan-European study (1990–2021), in 2021 the Russian Federation ranked among the top five countries by Quality of Care Index (QCI = 53.34), indicating a relatively favorable situation regarding ADR burden and pharmacovigilance performance. Additionally, data envelopment analysis (DEA), incorporating indicators such as the density of physicians, nurses, and pharmacists, also placed the Russian Federation among the five leading countries [10].

Pharmacogenomic testing continues to rely predominantly on the analysis of selected, well-characterized polymorphisms (e.g., in *CYP2C19, CYP2D6, VKORC1*) using PCR-based methods or genotyping arrays [11]. Despite the clinical implementation of next-generation sequencing (NGS) technologies and the availability of testing for key pharmacogenes, ADR-related mortality remains substantial (7.53% of all ADR outcomes in a South Korean study, 2019–2024 [12]), most commonly associated with anticoagulants, corticosteroids, and immunosuppressants. It should be noted that NGS has specific limitations. Genotyping of *CYP2D6* is technically challenging due to the presence of the *CYP2D7* pseudogene, gene conversion events, and copy number variations (CNVs). Moreover, *CYP2D6* exhibits marked polymorphism, as do *HLA* genes. Certain pharmacogenes demonstrate high sequence homology; for example, *CYP2C9* and *CYP2C19* share more than 90% sequence similarity. Additional complexity arises from multiallelic variants, such as *UGT1A1*28* (2-234668879-C-CAT), *UGT1A1*36* (2-234668879-CAT-C), and *UGT1A1*37* (2-234668879-C-CATAT), which require sequencing depths of at least 400× for reliable detection. In whole-exome sequencing (WES), further challenges stem from the lack of coverage of non-coding regions that may harbor clinically relevant variants, as exemplified by *VKORC1* [13–14]. These considerations often favor targeted NGS panels due to their optimal balance between cost, depth of coverage, and the ability to accurately detect structural variants (SVs) [15]. Nevertheless, within the framework of a single, accessible genetic test such as WES—which simultaneously enables the identification of causative variants underlying a patient’s disease as well as carrier status for pathogenic variants— pharmacogenomic testing (with the aforementioned analytical limitations) may serve as an effective screening approach.

In Russia, the economic consequences of ADRs and their assessment methods have become a subject of increasing interest. Studies indicate a substantial economic burden and underscore the need for preventive strategies [16]. However, large-scale population data on clinically relevant pharmacogenetic variants in the Russian Federation remain limited, and centralized pharmacogenetic testing has not yet been implemented [17, 18]. The aim of the present study was to analyze the frequencies of clinically significant pharmacogene variants and *HLA* alleles in a large population cohort (over 6,000 exomes generated between 2020 and 2025) and to assess their potential implications for drug efficacy, metabolism, dosing, and toxicity.

## Materials and Methods

### 2.1. Study Cohort

The study included 6,102 anonymized exome samples obtained between 2020 and 2025 from individuals of various ages (including neonates); 3,169 samples were from females and 2,933 from males. The cohort was heterogeneous and comprised both individuals without a confirmed diagnosis (including couples planning pregnancy and undergoing carrier screening for pathogenic variants in genes associated with hereditary diseases) and patients with suspected hereditary disorders (monogenic and digenic forms) evaluated in pediatric and adult inpatient settings. In some cases, parental samples were analyzed as part of segregation studies.

The majority of samples were obtained from medical institutions in Moscow; a subset of patients was hospitalized in federal centers, including the Russian Children’s Clinical Hospital, Petrovsky National Research Center of Surgery, Filatov Children’s City Clinical Hospital, Herzen Moscow Oncology Research Institute, Blokhin National Medical Research Center of Oncology, and Dmitry Rogachev National Medical Research Center of Pediatric Hematology, Oncology and Immunology.

For all samples, minimal demographic data (sex) and information on the type of exome enrichment kit were available.

### 2.2. WES

Genomic DNA (500 ng) extracted from peripheral blood samples was used for library construction with the MGIEasy Universal DNA Library Prep Set (MGI Tech, Shenzhen, China). Fragmentation of DNA was achieved by ultrasonication on a Covaris S-220 system (Covaris, Inc., Woburn, MA, USA), yielding fragments with a mean length of approximately 250 bp. Targeted exome capture was carried out with Agilent SureSelect Human All Exon enrichment kits (v6, v7, and v8) [19]. Quantification of DNA and prepared libraries was performed on a Qubit Flex fluorometer using the dsDNA HS Assay Kit (Invitrogen, Waltham, MA, USA). Library size distribution and integrity were evaluated with the Bioanalyzer 2100 platform (Agilent Technologies, Santa Clara, CA, USA). Following circularization, the libraries were sequenced in paired-end mode (PE100) on the DNBSEQ-G400 instrument (MGI Tech, Shenzhen, China), resulting in an average target coverage of approximately 100×.

FastQ files were generated using basecallLite software (ver. 1.0.7.84; MGI Tech, Shenzhen, China). Raw sequencing data quality was assessed using FastQC v0.11.9 (Babraham Institute, Cambridge, UK) [20]. Based on quality control results, read correction and filtering were performed with BBDuk v38.96 [21]. Reads were aligned to the human reference genome GRCh38 using BWA-MEM v2.2.1 [22], followed by alignment file processing with SAMtools v1.9 [23]. Read deduplication and exome enrichment quality metrics were calculated using Picard v2.22.4 [24].

Samples with inadequate WES quality metrics were excluded, including those with evidence of contamination, mean coverage below 60×, or target coverage completeness <95% at ≥10× depth. Across the cohort, the median on-target coverage was 100.35× and the mean coverage was 111×. The proportion of target nucleotides covered at ≥10×, ≥20×, and ≥30× was 97.2%, 95.6%, and 92.9%, respectively (Table S1).

Single nucleotide variants (SNVs) and short insertions/deletions (indels) were identified using a standard pipeline based on bcftools v1.18 [25]. To ensure compatibility with pharmacogenetic databases, VCF files were additionally annotated with rsIDs using dbSNP build 156 via bcftools annotate.

CNV analysis was performed using CNVkit v0.9.8 [26], and identified CNVs were annotated with ClassifyCNV v1.1.1 [27]. *HLA* alleles were determined using HLA-HD (ver. 1.7.0) [28], designed for high-resolution *HLA* typing from high-throughput sequencing data. Analysis was conducted on GRCh38-aligned BAM files. The resulting *HLA* alleles were converted to G-group format and subsequently integrated into the pharmacogenetic analysis.

### 2.3. Generation of Pharmacogenomic Reports

Only variants with confidently determined genotypes (excluding records with missing GT fields) were included in the analysis. Using PAnno [29], diplotypes were assigned—based on European population data—for *CYP2B6, CYP2C19, CYP2C8, CYP2C9, CYP2D6, CYP3A4, CYP3A5, CYP4F2, DPYD, NUDT15, SLCO1B1, TPMT, UGT1A1. CYP3A4∗36* (formerly *CYP3A4∗1G*), defined by NM_017460.6:c.1026+12G>A, was included in this study as it was part of the PAnno pipeline, despite its exclusion from current PharmVar definitions due to uncertain functional significance [30]. The results were subsequently integrated with *HLA* reports. Pharmacogenomic annotations were generated using PharmGKB (levels of evidence 1A–2B; version dated January 16, 2025) [31] and included information on genomic position, variant or haplotype, gene, level of evidence, associated drugs, phenotypes, allele functional significance, and the number of observations in the cohort.

The bioinformatics pipeline is schematically illustrated in Figure 1.

**Figure 1.**
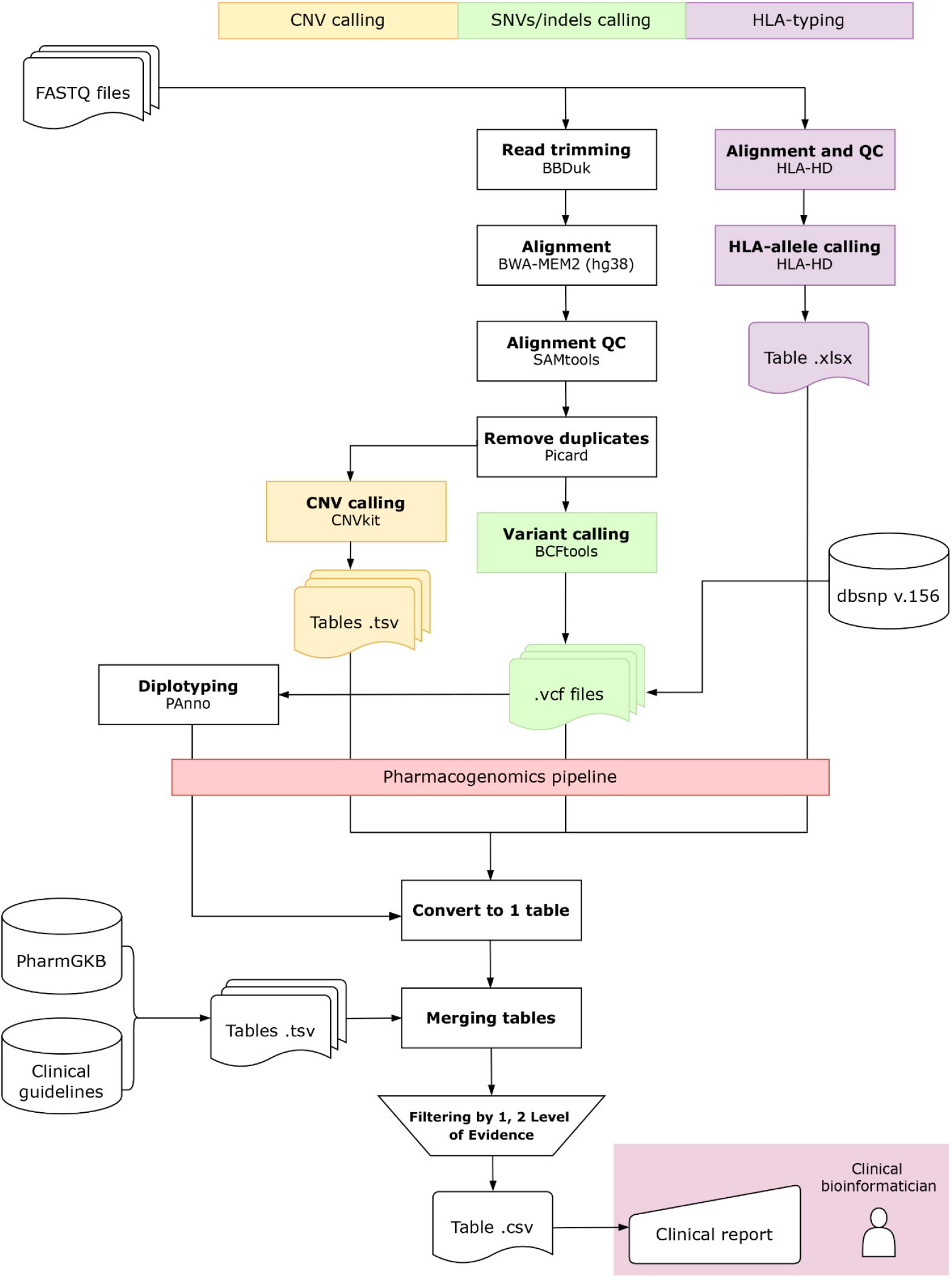
Bioinformatics pipeline for whole-exome sequencing (WES) data analysis used to generate pharmacogenomic and HLA reports, including quality control, alignment to the GRCh38 reference genome, detection of SNVs, indels, and CNVs, *HLA* allele typing, and pharmacogenetic variant annotation.

PharmVar [32] was used for allele analysis, and separate CNV reports were generated for 33 Very Important Pharmacogenes (VIPs) [31]. Metabolizer phenotype translation for *CYP2B6, CYP2C9, CYP2C19, CYP2D6* was based on the 2023 recommendations by Caudle KE et al. [33], whereas for the remaining genes, CPIC 2016 guidelines were applied [34].

## Results

### 3.1. Distribution of VIPs Alleles

Among *CYP2D6* alleles, the most frequent were *1 (35.45%), *2 (17.87%), *4 (16.35%), *35 (8.82%), and *41 (7.95%), overall consistent with distributions reported in European populations (Figure 2A, Table S2). In addition, *CYP2D6* exhibited the highest number of unique alleles—49 (Figure 2B). For *CYP2C19* the *1 allele predominated (78.27%), while *2 and *38 were observed at frequencies of 12.83% and 7.72%, respectively. Other decreased- or no-function variants were detected less frequently but collectively constituted a considerable proportion of individuals with potentially altered metabolic phenotypes (Figure 2C). In *CYP2C9*, the most common allele was *1 (81.39%), followed by *2 (9.47%) and *3 (7.85%), associated with decreased and absent enzymatic activity, respectively. For *SLCO1B1*, the frequencies of *1 and *15 were 53.33% and 17.09%, respectively. Both increased-function and no-function alleles in *SLCO1B1* accounted for a substantial proportion relative to other VIPs (14.25% and 20.73%, respectively). In *UGT1A1*, the most frequent alleles were *1 (62.81%) and *28 (33.75%). The compound allele *UGT1A1*80+*28* was not detected due to the location of *80 in a non-coding region. Decreased-function alleles in *UGT1A1* accounted for the highest proportion compared to other VIPs (35.96%). Analysis of *DPYD* using PAnno revealed predominance of the reference allele *1 (38.82%), followed by *9A (23%) and *5 (18.44%). Variants not assigned to star alleles comprised 14.35% (c.496A>G, c.1896T>C, c.775A>G, c.2846A>T, c.1774C>T, c.2161G>A, c.2279C>T, c.451A>G, c.934C>T). However, the PAnno algorithm failed to call c.1236G>A, as it is considered part of HapB3 together with c.1129-5923C>G (present in 3.87% of individuals in heterozygous state and 0.03% in homozygous state), and missed one heterozygous carrier of c.2303C>A; both were identified by independent SNV analysis. For *TPMT* and *NUDT15*, functional *1 alleles predominated (97.06% and 97.17%, respectively). In *CYP2B6*, the most frequent alleles were *1 (51.27%), *6 (21.35%), and *5 (11.84%). For *CYP3A4*, allele *1 was predominant (88.98%), whereas *36 was observed at a frequency of 9.56%. For *CYP3A5*, *1 was detected in nearly all cases (99.96%), which did not correspond to the expected distribution for the Russian population and was attributed to WES limitations (see Section 3.4). For *CYP4F2*, the most common allele was *1 (61.94%), followed by *3 (25.48%), associated with altered warfarin dose requirements. *CYP4F2*2* was observed at a frequency of 12.59%.

**Figure 2.**
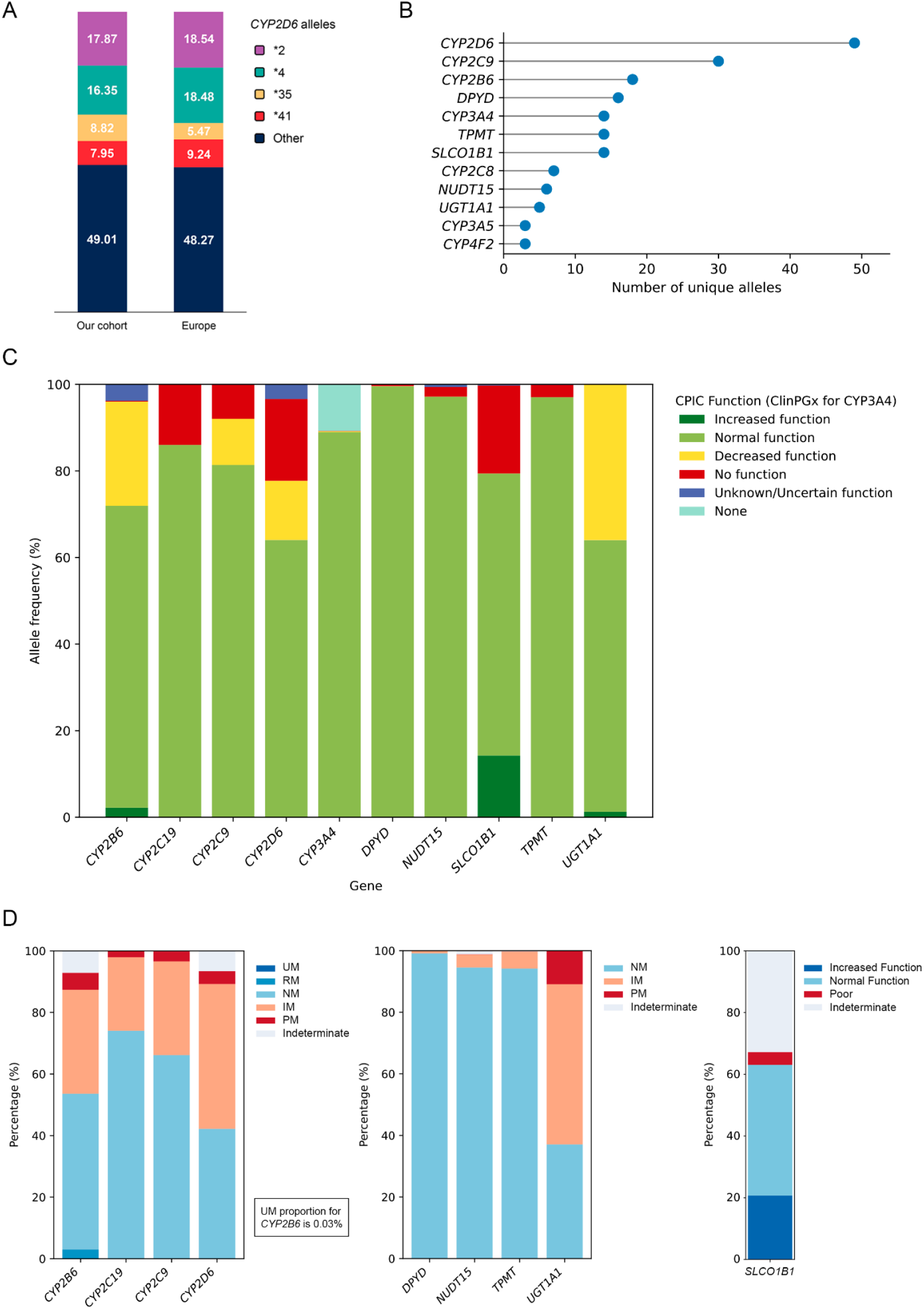
**A**. Percentage distribution of the most common *CYP2D6* (*2, *4, *35, *41, and others; *n* = 49) among study participants, consistent with European distributions. **B**. Number of unique alleles identified for each diplotyped pharmacogene (*CYP2B6, CYP2C19, CYP2C8, CYP2C9, CYP2D6, CYP3A4, CYP3A5, CYP4F2, DPYD, NUDT15, SLCO1B1, TPMT, UGT1A1*), illustrating high polymorphism at certain loci. **C**. Percentage distribution of alleles in *CYP2C19, CYP2C9, SLCO1B1, UGT1A1, DPYD, TPMT, NUDT15, CYP2B6, CYP3A4*, and *CYP2D6* according to functional classification (e.g., decreased or no function). **D**. Percentage distribution of metabolic phenotypes for key genes *CYP2B6, CYP2C19, CYP2C9, CYP2D6, DPYD, NUDT15, TPMT*, and *SLCO1B1*. Notes: Common non-normal alleles *CYP2C19*17* and *UGT1A1*80* were not called; therefore, interpretation should be approached with caution, as they were likely included within *1 alleles. Abbreviations: NM—Normal metabolizer, IM— Intermediate metabolizer, PM—Poor metabolizer, RM—Rapid metabolizer, UM—Ultrarapid metabolizer.

Among CYP family genes, the most frequent normal diplotypes were: *CYP3A4*1/*1* in 4,837 individuals (79.27%), *CYP2C8*1/*1* in 4,569 (74.88%), *CYP2C9*1/*1* in 4,036 (66.14%), *CYP2C19*1/*1* in 3,740 (61.29%), *CYP4F2*1/*1* in 3,388 (55.52%). For *UGT1A1*, the most common diplotype was *1/*28, detected in 2,900 individuals (47.53%), whereas *1/*1 was observed in 2,251 (36.89%). The homozygous *28/*28 diplotype was identified in 569 individuals (9.32%). In *SLCO1B1*, *1/*1 was detected in 1,723 individuals (28.24%), *1/*15 in 1,143 (18.73%), *15/*15 in 177 (2.9%), and *5/*15 in 71 (1.16%). For *CYP2B6*, the most common diplotype was *1/*1 (1,666 individuals; 27.3%), followed by *1/*6 (20.63%) and *1/*5 (12.47%), reflecting the high polymorphism of this gene. *CYP2D6* exhibited marked diplotype diversity. The most frequent was *1/*1 (876 individuals; 14.36%), followed by *1/*4 (11.5%), *1/*2 (11.47%), and *4/*4 (3.18%). For *DPYD*, the most common diplotype was *1/*1 (*n* = 1,350; 22.12%).

Notably, in 14 samples (0.23% of the cohort), PAnno generated multiple alternative diplotype assignments for *CYP2C19* (12 samples simultaneously assigned *CYP2C19*1, CYP2C19*2, CYP2C19*2, CYP2C19*11*), and *CYP2D6* (2 samples assigned *CYP2D6*4, CYP2D6*22, CYP2D6*4, CYP2D6*142*). In such cases, manual validation was performed based on primary VCF variant analysis using PharmVar coordinates and SNVs. Final assignments were revised to *CYP2C19*1*/*CYP2C19*2* and *CYP2D6*4*/*CYP2D6*142*.

Most participants exhibited normal (NM) or intermediate (IM) metabolizer phenotypes for the studied cytochrome P450 genes (Figure 2D). For *CYP2B6*, NM accounted for 50.59% and IM for 33.74%, whereas poor metabolizer (PM), rapid metabolizer (RM), and ultrarapid metabolizer (UM) were much less frequent (5.54%, 3%, and 0.03%, respectively). A similar distribution was observed for *CYP2C19* and *CYP2C9*, where NM predominated (74.06% and 66.14%, respectively), and PM did not exceed 3.3%. For *CYP2D6*, the distribution was shifted toward intermediate metabolism: IM was observed in 47% of individuals, NM in 42.22%, and PM in 4.23%. UM phenotypes for *CYP2D6* and *CYP2C19* were not detected in this cohort.

For *DPYD*, nearly all individuals were NM (99.15%), with IM comprising 0.85% (*n* = 52); PM was not observed. For *NUDT15* and *TPMT*, the vast majority were NM (94.54% and 94.18%, respectively), with very low PM frequency (0.06% for both). In contrast, PM phenotype for *UGT1A1* was present in 10.9% of individuals, while NM accounted for only 37.09%. For *SLCO1B1*, a more complex classification was applied: 42.31% were classified as NM, 32.79% as indeterminate, 20.75% as increased function, and 4.15% as poor function, reflecting the presence of alleles with uncertain or insufficiently characterized functional significance.

### 3.2. Pharmacogenetically Relevant HLA Alleles

Analysis of *HLA* allele frequencies demonstrated high polymorphism across both class I (*HLA-A, HLA-B, HLA-C*) and class II (*HLA-DPB1, HLA-DQA1, HLA-DQB1, HLA-DRB1*) genes. Among *HLA* class I genes, the most frequent *HLA-A* alleles were *HLA-A*02* (29.42%), including *HLA-A*02:01* (26.12%) followed by *HLA-A*03* (13.52%) and *HLA-A*01* (11.97%). For *HLA-C*, the predominant alleles were *HLA-C*07* (27.03%), *HLA-C*04* (13.25%), and *HLA-C*06* (12.55%). The *HLA-B* locus showed a relatively even distribution, with *HLA-B*07* (11.68%), *HLA-B*35* (11.06%), *HLA-B*44* (9.21%), and *HLA-B*18* (6.92%) being the most common.

*HLA* class II genes also exhibited several high-frequency alleles. For *HLA-DPB1*, the most prevalent allele was *HLA-DPB1*04* (54.91%), including *HLA-DPB1*04:01* (40.45%), markedly exceeding other variants. At the *HLA-DQA1* locus, *HLA-DQA1*01* (42.95%) and *HLA-DQA1*05* (27.02%) predominated. For *HLA-DQB1*, the most frequent alleles were *HLA-DQB1*03* (34.46%), *HLA-DQB1*06* (22.89%), and *HLA-DQB1*05* (20.11%). In *HLA-DRB1*, several alleles displayed comparable frequencies, including *HLA-DRB1*07* (13.91%), *HLA-DRB1*11* (13.62%), *HLA-DRB1*15* (13.33%), and *HLA-DRB1*13* (13.11%), reflecting substantial allelic heterogeneity at this locus (Table S3).

### 3.3 Overview of Pharmacogenomic Annotations

Within PharmGKB level 1A–2B annotations, 663 clinically significant annotations were identified, including 56 related to dosing, 65 to efficacy, 334 to metabolism, 196 to toxicity, and 12 belonging to other clinical categories (Table S4; Figure 3A).

**Figure 3.**
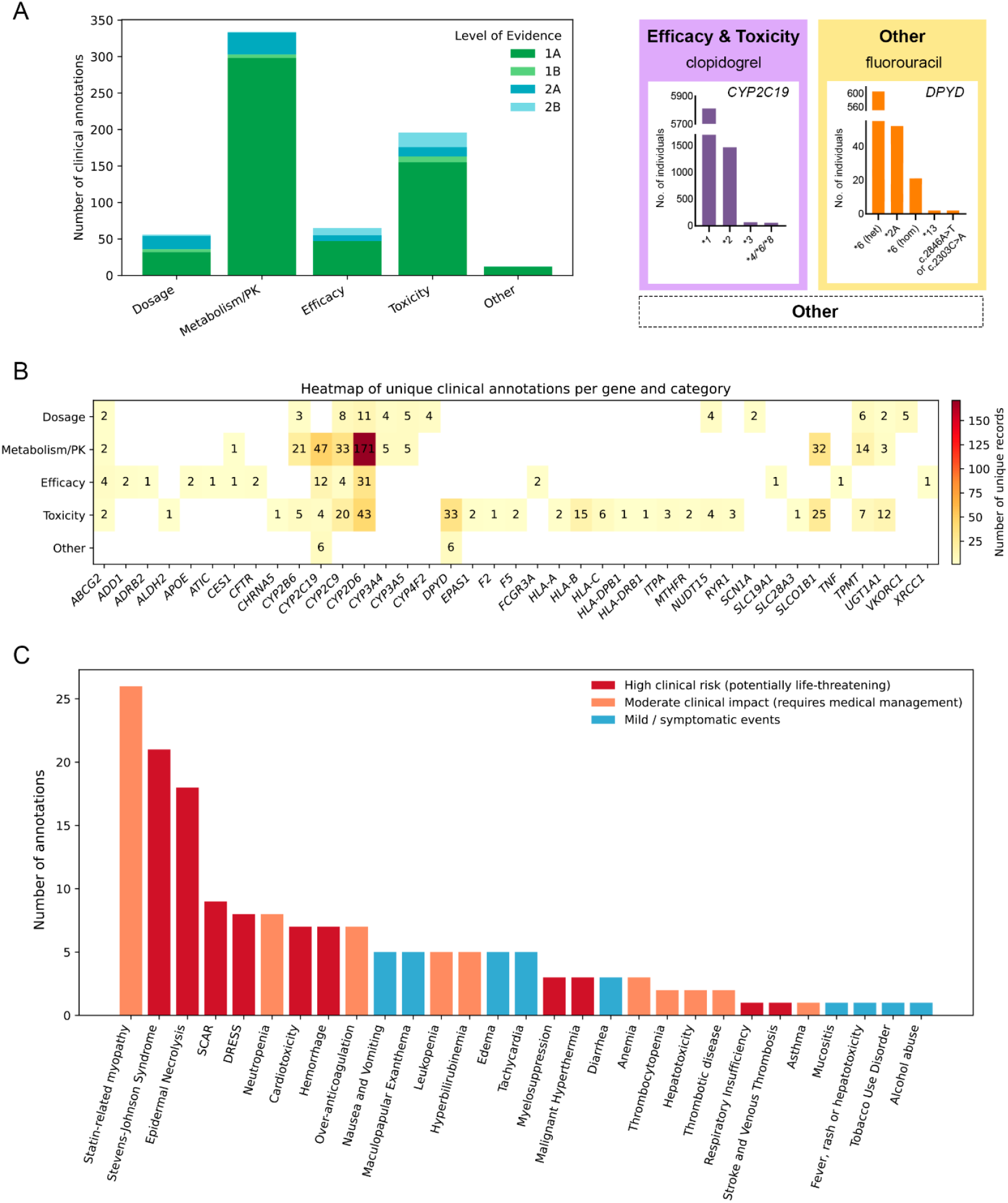
**A**. Clinically significant PharmGKB annotations (levels 1A–2B) distributed by category: dosing, efficacy, metabolism/pharmacokinetics, toxicity, and other. **B**. Number of unique clinical annotations per gene (levels 1A–2B). The highest number of records was associated with *CYP2D6*. **C**. Expected adverse effects. Hepatotoxicity—Drug-induced liver injury or Toxic liver disease. Abbreviations: SCAR—Severe Cutaneous Adverse Reactions, DRESS—Drug Reaction with Eosinophilia and Systemic Symptoms.

The highest number of dose-adjustment recommendations was associated with *CYP2D6* (*n* = 11, including 2 related to *CYP2D6*5*) and *CYP2C9* (*n* = 8), followed by *TPMT* (*n* = 6), *CYP3A5* (*n* = 5), and *VKORC1* (*n* = 5) (Figure 3B). These genes were linked to anticoagulants (acenocoumarol, phenprocoumon, warfarin), immunosuppressants and thiopurines (azathioprine, mercaptopurine, thioguanine, tacrolimus), the antiepileptic drug carbamazepine, the tricyclic antidepressant imipramine, the antineoplastic agent irinotecan, the antiretroviral drug efavirenz, opioid analgesics (fentanyl, tramadol), the β-adrenergic blocker metoprolol, and allopurinol, which affects uric acid metabolism.

The “metabolism” category was the most represented and included 334 annotations, nearly half of which were associated with *CYP2D6* (*n* = 171, including 24 related to *CYP2D6*5*). Substantial contributions were also observed for *CYP2C19* (*n* = 47), *CYP2C9* (*n* = 33), *SLCO1B1* (*n* = 32), and *CYP2B6* (n = 21). These findings underscore the dominant role of variability in biotransformation enzymes in interindividual differences in the pharmacokinetics of a wide range of drugs, including psychotropic agents (*n* = 22), nonsteroidal anti-inflammatory drugs (celecoxib, flurbiprofen, ibuprofen, lornoxicam, meloxicam, piroxicam, tenoxicam), statins (atorvastatin, fluvastatin, lovastatin/lovastatin acid, pitavastatin, pravastatin, rosuvastatin, simvastatin/simvastatin acid), opioid analgesics (codeine, hydrocodone, methadone, oxycodone, tramadol), proton pump inhibitors (dexlansoprazole, lansoprazole, omeprazole, pantoprazole, rabeprazole), immunosuppressants and thiopurines (azathioprine, mercaptopurine, thioguanine, tacrolimus), antiarrhythmic agents (phenytoin, flecainide, propafenone), antineoplastic agents (tamoxifen, SN-38), antiretroviral drugs (efavirenz, nevirapine), the antiplatelet agent clopidogrel, the antiemetic tropisetron, the antifungal agent voriconazole, and the immunomodulator siponimod.

Annotations related to therapeutic efficacy (*n* = 60) were primarily associated with *CYP2D6* (*n* = 31, including 5 related to *CYP2D6*5*) and *CYP2C19* (*n* = 12), but also included pharmacodynamic and transporter genes such as *ABCG2* (*n* = 4) and *FCGR3A* (*n* = 2), reflecting the contribution of both metabolic and receptor-mediated mechanisms to therapeutic response. These annotations encompassed antineoplastic and hormonal therapies (tamoxifen, platinum compounds), anticoagulants and antiplatelet agents (clopidogrel, warfarin), opioid analgesics (codeine, hydrocodone, tramadol), statins (atorvastatin, rosuvastatin), proton pump inhibitors (lansoprazole, omeprazole, pantoprazole), immunosuppressive and targeted biological agents (methotrexate, etanercept, rituximab), antiemetics (ondansetron, tropisetron), the diuretic hydrochlorothiazide, the β_2_-agonist salmeterol, allopurinol, and the cystic fibrosis therapy ivacaftor.

The “toxicity” category comprised 196 annotations and was characterized by heterogeneous contributions from metabolic and immunogenetic factors. The largest number of annotations was associated with *CYP2D6* (*n* = 43, including 7 related to *CYP2D6*5*), *DPYD* (*n* = 33), *SLCO1B1* (*n* = 25), and *CYP2C9* (*n* = 20). A substantial contribution of *HLA-B* (*n* = 15) highlighted the role of HLA alleles in immune-mediated ADRs. These associations were considered broadly relevant to therapy, in contrast to dosing recommendations adjusted for age, sex, body weight, and other individual characteristics. The implicated drugs included antiretroviral agents (abacavir, atazanavir/ritonavir, efavirenz, nevirapine), anticoagulants and antiplatelet agents (acenocoumarol, warfarin, aspirin), antidepressants (amitriptyline, clomipramine, desipramine, imipramine, nortriptyline, venlafaxine, citalopram), cytotoxic and antimetabolite anticancer agents (anthracyclines and related compounds, daunorubicin, doxorubicin, capecitabine, fluorouracil, tegafur, FOLFIRI, irinotecan, sorafenib), immunosuppressive and immunomodulatory agents (azathioprine, methotrexate, mercaptopurine, thioguanine, peginterferon alfa-2b with ribavirin), antiepileptic drugs (carbamazepine, lamotrigine, oxcarbazepine, phenytoin), opioid and centrally acting analgesics (codeine, tramadol), statins and HMG-CoA reductase inhibitors (atorvastatin, fluvastatin, lovastatin, pravastatin, rosuvastatin, simvastatin), antibacterial and antimicrobial agents (dapsone, flucloxacillin, sulfamethoxazole/trimethoprim), inhalational anesthetics and neuromuscular blocking agents (desflurane, enflurane, halothane, isoflurane, methoxyflurane, sevoflurane, succinylcholine), systemic hormonal contraceptives, psychoactive substances and stimulants (atomoxetine, nicotine, ethanol), allopurinol, and the antiglaucoma drug methazolamide.

The most frequently annotated ADR was statin-associated myopathy (*n* = 26) (Figure 3C). Severe cutaneous adverse reactions (SCAR) included Stevens–Johnson syndrome (*n* = 21), toxic epidermal necrolysis (*n* = 18), and DRESS syndrome (*n* = 8); an additional 9 cases were annotated as SCAR without further specification. Hematological complications included neutropenia (*n* = 8), leukopenia (*n* = 5), anemia (*n* = 3), myelosuppression (*n* = 3), and thrombocytopenia (*n* = 2). Hemostatic disturbances included excessive anticoagulation (*n* = 7), hemorrhagic complications (*n* = 7), and thrombotic events, including thrombotic disease and stroke with venous thrombosis (4 individuals in total). Cardiotoxic effects were annotated in 7 individuals and included tachycardia (*n* = 5) and edema (*n* = 5). Hepatic and gastrointestinal manifestations included hyperbilirubinemia (*n* = 5), nausea and vomiting (*n* = 5), diarrhea (*n* = 3), drug-induced liver injury (*n* = 2), and mucositis (*n* = 1). Less frequently reported were potentially life-threatening conditions such as malignant hyperthermia (*n* = 3), respiratory failure (*n* = 1), and exacerbation of bronchial asthma (*n* = 1). Some annotations reflected associations with behavioral phenotypes, including tobacco use disorder and alcohol-related disorders (1 individual each). It is critically important to emphasize that these numbers represent the count of individuals for whom a PharmGKB annotation indicated a potential association between a genetic variant and a given ADR, rather than the actual observed incidence of the clinical event. Moreover, the presence of such an annotation does not imply an obligatorily increased risk for a specific individual, as interpretation should be based on the diplotype and corresponding metabolizer phenotype rather than on the presence of a single allele. In some cases, similar annotations are also present for normal-function alleles, limiting direct extrapolation to individual risk.

The “other” category (*n* = 12) was predominantly represented by variants in *CYP2C19* (*n* = 6) and *DPYD* (*n* = 6) and included clinical effects not directly related to dosing, efficacy, or toxicity (limited to clopidogrel and fluorouracil) (Figure 3A).

It should be noted that several annotations are associated with variants located in non-coding regions, including *CYP3A5*3* and *CYP2C19*17*, the detection of which in the present study was limited by targeted enrichment design and annotation algorithms. Therefore, interpretation of the clinical relevance of these alleles, as well as other variants outside coding regions, requires caution and is addressed separately (see Section 3.4).

### 3.4 Limitations of WES Data

Analysis of WES data demonstrated that a proportion of clinically significant pharmacogenetic variants are located in non-coding regions of the genome and, consequently, are either completely absent from WES datasets or characterized by insufficient and uneven coverage. This primarily concerns variants in promoter regions, introns, and 3′ untranslated regions (3′UTRs).

Specifically, for *CYP3A5*, the clinically significant variant rs776746 (c.253-1A>G, allele *3), which determines enzyme expression and is critically important for tacrolimus therapy interpretation, is located in a region with inadequate coverage. In all analyzed samples, the exonic variant rs28371764 (*1C) was successfully called, leading annotation pipelines to automatically assign the homozygous *1/*1 genotype. Thus, the absence of data for rs776746 was interpreted algorithmically as absence of the alternative allele rather than genotype uncertainty, representing a systematic limitation of the exome-based approach.

Similar limitations were identified for non-coding variants in *CYP2B6* (rs34223104, c.-82T>C, alleles *22, *34, *35, *36), *CYP2C19* (rs12248560, c.-806C>T, alleles *4, *17), *DPYD* (rs75017182, c.1129-5923C>G), *CYP3A4* (rs35599367, c.522-191C>T, allele *22), and *UGT1A1* (rs887829, -c.364C>T, allele *80; as well as the CA(TA)_n_TAA, repeat corresponding to allele *28). However, the non-coding variant *CYP2C19* rs12769205 (c.332-23A>T, alleles *2, *35) was located in a region of stable exome coverage >10× and was detected on 1,582 chromosomes (12.96%). Similarly, *CYP2B6*22* (156 alleles, 1.28%) and *UGT1A1*\*28 (9.32% homozygotes) were reliably called. In addition, PAnno identified *CYP2C19*4* on 21 chromosomes (0.17%), but not *CYP2C19*17*.

For the pharmacogenes *CYP2C8, CYP2C9, CYP2A13, CYP4F2, NAT2, NUDT15, SLCO1B1*, and *TPMT* no clinically significant non-coding variants with a level of evidence ≥2B were identified; accordingly, no exome coverage-related limitations were observed for these genes.

Additional analysis of deep intronic and intergenic variants present in the PharmGKB database (January 2025) with evidence levels up to 2B demonstrated that all such variants were characterized by low or absent coverage in WES. This applied to rs12777823 (*CTBP2P2*), rs2359612 and rs9923231 (*VKORC1*), rs8099917 (*IFNL3*), rs10929302 (*UGT1A6*) and rs1800629 (*TNF*). Visual inspection using IGV confirmed a systematic absence of informative reads in the corresponding regions.

Collectively, these limitations preclude accurate pharmacogenetic interpretation for at least 20 drugs, including warfarin and other vitamin K antagonists, tacrolimus, fluoropyrimidines, proton pump inhibitors, selective serotonin reuptake inhibitors and tricyclic antidepressants, as well as interferon-based therapies.

Comparison of WES data obtained from the same sample using different versions of exome enrichment kits (SureSelect Human All Exon v6, v7, and v8) revealed rare but clinically significant discrepancies in *HLA* alleles and VIP diplotype calling (Doc S1). These differences are likely attributable to changes in probe design and target composition (e.g., intersection of v7 and v8 bed files = 34.84 Mb, according to our previous study [35]), and are supported by minor differences in SNV and indel calling metrics (within intersected targets, v8 detected +3.06% SNVs and +8.49% indels). For v6–v7 comparisons, discrepancies were observed only in *UGT1A1* (8 individuals, *1 and *28), with no differences in *HLA* alleles. In v7–v8 comparisons, discrepancies were more heterogeneous: *UGT1A1* – 9 individuals (*1 and *28), *SLCO1B1* – 2 individuals (*37 and *15, 1* and *14), *HLA* – 4 individuals, one discrepancy per individual (*HLA-C*06:02* −> *HLA-C*18:01, HLA-C*07:02* −> *HLA-C*07:27, HLA-B*13:01* −> *HLA-B*13:02, HLA-DQA1*02:01* −> *HLA-DQA1*05:01*). These discrepancies may affect pharmacogenetic interpretation of therapies involving irinotecan, statins, and immunomodulators, particularly with respect to dosing and toxicity risk.

In WES data, *CYP2D6*1xN*, *2xN, *4xN, and *35xN are difficult to detect due to CNV, hybrid alleles involving closely related pseudogenes *CYP2D7* and *CYP2D8*, and intrinsic methodological limitations. *CYP2D6*61* and *CYP2D6*63* represent hybrid *CYP2D6::CYP2D7* gene associated with reduced and absent CYP2D6 enzymatic activity, respectively, in the metabolism of N-desmethyltamoxifen compared *CYP2D6*1* (Level 3 evidence). Consequently, definitive therapeutic recommendations cannot be reliably established for certain opioid analgesics (codeine, oxycodone, tramadol), antidepressants (desipramine, nortriptyline, paroxetine, mirtazapine, doxepin), antiemetics (ondansetron, tropisetron), and psychotropic agents (risperidone, atomoxetine) (total *n* = 12) [36]. Additional limitations exist for *CYP2B6*30*, a SV representing a *CYP2B6::CYP2B7* hybrid gene. Individuals carrying this allele in combination with a normal-function allele may demonstrate reduced bupropion metabolism compared with carriers of two normal-function alleles (Level 2B evidence). *CYP2A6*4* corresponds to a complete gene deletion, resulting in absence of enzyme activity, whereas *CYP2A6*1×2* represents duplication of a functional allele and may increase substrate metabolism. For both variants, high-level clinical evidence (>Level 2B) is available only for nicotine metabolism.

### 3.5 Assessment of the Feasibility of Imputing Non-Coding Pharmacogenetic Variants from WES Data

To evaluate the theoretical feasibility of reconstructing non-coding pharmacogenetic variants from WES data, a prototype imputation file based on linkage disequilibrium (LD) analysis was developed and tested. The reference panel consisted of 814 whole genomes, including 229 samples from the V. P. Serbsky National Medical Research Center of Psychiatry and Narcology and 585 samples from the Russian Gerontology Research and Clinical Center (bioinformatic analysis as described in Section 2.2). LD coefficients (r^2^) were calculated using PLINK v1.90b6.24 (--r2) [37] for all clinically significant non-coding variants identified in the previous analysis stage.

The results demonstrated that for the target variants, no exonic or capture-covered non-exonic markers in strong LD were identified. Specifically, clinically significant variants *VKORC1* rs9923231 (c.-1639G>A), *TNF* rs1800629, *CYP3A5* rs776746 (c.253-1A>G), and *CYP2C19* rs12248560 (c.-806C>T) were entirely absent from LD reports involving variants potentially covered by WES. This finding is particularly important for rs776746 (*CYP3A5*3*), given its high population frequency in the Russian population.

For several other variants, LD was observed exclusively with other non-coding markers, which does not resolve the problem of genotype reconstruction using WES. For example, *DPYD* c.1129-5923C>G (rs75017182) demonstrated moderate LD with intronic rs2811211 (r^2^ = 0.49), which in turn was linked to another deep intronic variant rs146353874 (r^2^ = 0.477); however, none of these markers were associated with exonic variants. A similar pattern was observed for rs12777823 (*CTBP2P2*), where LD with rs12772169 (r^2^ = 0.559) was not accompanied by linkage to variants captured by WES.

The variant rs8099917 (*IFNL3*) was also unsuitable for WES-based imputation: although it showed strong LD with several neighboring markers, all were located several kilobases from the nearest exon boundaries and were therefore absent from WES data. Likewise, rs10929302 (*UGT1A6*) demonstrated only moderate LD with neighboring non-coding variants (r^2^ < 0.6), precluding their use as reliable proxies.

The only variant demonstrating a potential for limited imputation was rs2359612 (*VKORC1*), which showed moderate LD with several neighboring markers: with chr16:31092871:C:CA (r^2^ = 0.355); this variant was in turn associated with rs8050894 (r^2^ = 0.3738), which further demonstrated stronger linkage with rs9934438 (r^2^ = 0.693; Level of Evidence 3 for warfarin). However, even in this case, LD values remained below the commonly accepted imputation threshold (r^2^ = 0.8), limiting the practical utility of this approach. In the Russian Federation, the prevalence of rs2359612 is 59% and rs9934438 is 41% according to the CSP FMBA database [38]; however, rs9923231 is detected at nearly the same frequency (41%), differing by only 0.114%.

Particular attention should be given to the *UGT1A1* variant cluster, where alleles *28 and *80 demonstrated strong LD (r^2^ = 0.984). In this case, however, the practical value of imputation was limited because the clinically significant *28 allele itself is covered by WES and can be directly determined, rendering imputation of the functionally uncertain *80 allele redundant.

## Discussion

In the Russian population, the prevalence of alleles in genes of the cytochrome P450 family varies, with several alleles exceeding frequency thresholds of 0.5–1% and thus having practical relevance for pharmacogenetic testing. These include *CYP2C9* (*1, *2, *3, *29), *CYP2C19* (*1, *2, *3, *17), and *CYP2D6* (*1, *2, *3, *4, *6, *9, *10, *33, *35, *39, *41, *59) [17]. In the present study, analysis of the *CYP2C19*17* allele was limited because the key diagnostic variant c.-806C>T was absent from the target enrichment panel. As a result, PAnno automatically classified the corresponding haplotypes as *CYP2C19*1* (allele frequency in our cohort is 78.27%), since the shared variant p.I331 was present and no additional defining variant for alternative alleles was detected. According to Yudin V.S. et al. [17], the frequencies of *CYP2C19*1* and *17 in Russia are 52.99% and 25.55%, respectively, underscoring the need for careful interpretation of WES-based data (approximately one in four individuals would otherwise receive an incorrect diplotype assignment).

The frequencies of *CYP2C9*1*, *2, *3, and *29 were 81.39%, 9.47%, 7.85%, and 0.93%, respectively, with deviations from the CSP FMBA reference data of less than 0.85 percentage points for each allele. The cumulative absolute deviation across the four most common alleles did not exceed 1.1%, indicating high reproducibility of WES data for *CYP2C9*, likely due to the absence of clinically relevant non-coding variants in pharmacogenomic panels for this gene. For *CYP2D6*, the rank order of the most prevalent alleles fully matched reference data: *CYP2D6*1* (35.45%), *2 (17.89%), *4 (16.35%), *35 (8.82%), and *41 (7.95%). However, the difference in *CYP2D6*1* frequency compared with CSP FMBA data exceeded 2% (37.86%). A plausible explanation is the presence of at least eight star alleles containing the non-coding variant c.985+39G>T, which may occasionally escape variant calling, thereby altering allele distribution estimates.

Concordance between our cohort and European populations [31] was observed for the most frequent alleles of *CYP2B6* (*1, *6, *5, and *2), *DPYD* (*9A, *5, c.496A>G, *6, and c.1896T>C; the next most frequent European haplotype, HapB3 at 2.37%, was not called in WES), as well as complete agreement in both rank order and number of alleles for *CYP4F2*. For *TPMT*, concordance was also observed for common alleles *1, *3, and 2; however, *TPMT*12* detected in our cohort at 2.46% was not observed in Europeans according to the “TPMT Frequency Table” in PharmGKB (gnomAD v4.1.0 NFE frequency: 0.04691%). In contrast, *SLCO1B1*15* and *37 were detected in 17.09% and 11.85% of our cohort, respectively, compared with 15.02% and 25.27% in Europeans. Similarly, the relative frequencies of *NUDT15*3* and *6 were reversed (2.15% and 0.54% in our cohort vs. 0.2% and 0.3% in Europeans). *CYP3A4∗36* was present in 9.56% of our cohort, comparable to 9.03% among NFE in gnomAD. *CYP3A5∗6* and *CYP3A5∗7* were detected at 0.025% and 0.016%, respectively (vs. 0.072% and 0.0062% in NFE). The *UGT1A1*80* was not detected and was incorrectly classified as *UGT1A1*1*; in Europeans, *80 is in near-complete LD with *28 and has a frequency of 31.42%. Consequently, the observed frequency of *UGT1A1*1* in our cohort was inflated to 62.81%.

It should be noted that three genes were not diplotyped: *CYP1A2* (only Level 3 evidence for clozapine), *NAT2* (associated solely with isoniazid therapy for tuberculosis), and *CYP2A6* (associated with nicotine). Nevertheless, rsID-based annotations exist for these genes and should theoretically have been detectable according to our bioinformatic pipeline. Furthermore, the development of a universal tool for imputing clinically significant non-coding pharmacogenetic variants from WES data is currently not feasible for the Russian population.

The contribution of immunogenetic factors to ADRs is supported by the high frequency of clinically relevant *HLA* alleles in the studied cohort. The three most prevalent pharmacogenetically annotated alleles—*HLA-C*04:01* (13%), *HLA-C*06:02* (12.19%), and *HLA-DPB1*03:01* (10.77%)—are associated with life-threatening conditions such as Stevens– Johnson syndrome, toxic epidermal necrolysis, and aspirin-induced asthma. This indicates a substantial population-level risk of severe complications upon exposure to the corresponding drugs (nevirapine, sulfamethoxazole/trimethoprim, aspirin). Importantly, for carbamazepine prescribed in epilepsy, a clinically significant frequency (2.56%) *HLA-A*31:01* was observed, including annotations relevant to pediatric patients. Given the global prevalence of epilepsy of approximately 0.7% as of 2021 [39], this finding may be particularly important for preventing DRESS syndrome, especially in residents of the Perm region, the Republic of Kalmykia, Buryatia, Tuva, and Sakha [18].

A 2021 systematic review encompassing 108 studies across 39 drugs demonstrated that pharmacogenomic testing was cost-effective or cost-saving in 71% of cases [40]. In our cohort, alleles associated with reduced response to standard therapies exhibited high population frequencies (>15%). For example, 24.04% of individuals carried *CYP2C19*2*, potentially resulting in inadequate antiplatelet response to clopidogrel. *CYP2C9*2* and *3 were present in 18.01% and 15.06% of individuals, respectively, complicating warfarin dose optimization. A substantial proportion of the population (29.51% carriers of *CYP2D6*4*, while 3.18% of all individuals were homozygous for this allele) may experience reduced analgesic response to codeine and tramadol or poorer outcomes in tamoxifen-treated breast cancer. Prescribing such medications to carriers of reduced-function alleles without prior testing leads to direct economic losses due to lack of therapeutic effect, repeated consultations, hospitalizations, and treatment modification.

Implementation of these findings in clinical practice in the Russian Federation faces not only technical constraints but also regulatory barriers. Interpretation of pharmacogenetic results and recommendation of alternative therapy fall exclusively within the competence of a clinical pharmacologist, whereas a medical geneticist is authorized to prescribe only pathogenetic therapy (i.e., cannot prescribe analgesics or antibiotics), and pharmacogenomics is not part of routine genetic practice. Currently, Russian hospitals are not mandated to perform NGS-based pharmacogene analysis or even minimal testing of common *CYP2C19* and *CYP2D6*. Thus, our study addresses a fundamental objective—establishing a large-scale population database of pharmacogenetically relevant variant frequencies, which constitutes an essential scientific foundation for developing clinical guidelines, educational programs for specialists, and ultimately integrating personalized pharmacotherapy into routine care.

## Conclusions

This study presents a comprehensive analysis of clinically significant pharmacogenetic variants, *HLA* alleles, and CNVs in a large cohort of 6,102 exome samples. The findings characterize the contemporary structure of variability in key pharmacogenes within the Russian population and demonstrate the contribution of both metabolic and immunogenetic factors to interindividual variability in drug response.

Nevertheless, diplotype interpretation for certain genes should be approached with caution. The generated frequency estimates may serve, at least partially, as a reference framework for the development and adaptation of clinical pharmacogenetic guidelines, ADR risk assessment, and justification for implementing personalized pharmacotherapy. At the same time, the results underscore the inherent limitations of WES and the need for alternative methodological approaches when making therapeutic decisions.

## Supporting information

...with distributions reported in European populations (Figure 2A, Table S2).

...HLA-DRB1*15 (13.33%), and HLA-DRB1*13 (13.11%), reflecting substantial allelic heterogeneity at this locus (Table S3).

...and 12 belonging to other clinical categories (Table S4; Figure 3A).

## Supplementary Material

Table S1. Sequencing quality metrics. Table S2. Pharmacogenic Allele Frequency. Table S3. *HLA* Allele Frequency. Table S4. Summary of pharmacogenetic annotations. Doc S1. Comparison of exome enrichment kits in the context of NGS-based pharmacogenomic interpretation (v6, v7, v8).

## Institutional Review Board Statement

This study was conducted in accordance with the Declaration of Helsinki, and approval was obtained from the Local Research Ethics Committee of Russian National Medical University (Protocol No. 241, from 26 June 2024), and all participants provided written informed consent prior to data collection.

## Author Contributions

Buianova A.A. – formal analysis, conceptualization, methodology, software, writing–original draft preparation; Cheranev V.V., Kuznetsov M.Iu. – data curation, software; Shmitko A.O., Ilyina G.A., Belova V.A. – investigation; Vasiliadis Iu.A., Suchalko O.N. – software; Korostin D.O. – resources, supervision, writing–review and editing.

## Informed Consent Statement

Informed consent was obtained from all subjects involved in this study.

## Data Availability Statement

The sequence data are generated from patient samples and therefore are only available under restricted access.

